# A novel decision-making tool for first-line treatment selection in metastatic non-small cell lung cancer based on plasma proteome profiling

**DOI:** 10.1101/2022.12.01.22282769

**Authors:** Petros Christopoulos, Michal Harel, Coren Lahav, Itamar Sela, Nili Dahan, Niels Reinmuth, Ina Koch, Alona Zer, Mor Moskovitz, Adva Levy-Barda, Michal Lotem, Hovav Nechushtan, Rivka Katzenelson, Abed Agbarya, Mahmoud Abu-Amna, Maya Gottfried, Ella Tepper, Christine B. Ambrosone, Ido Wolf, Yanyan Lou, Raya Leibowitz, Adam P. Dicker, David P. Carbone, David Gandara

## Abstract

**Importance:** Advanced stage non-small cell lung cancer (NSCLC) patients with no driver mutations are typically treated with immune checkpoint inhibitor (ICI)-based therapy, either in the form of monotherapy or concurrently with chemotherapy, while treatment modality selection is based mainly on programmed death ligand 1 (PD-L1) expression levels in the tumor. However, PD-L1 assays are only moderately predictive of therapeutic benefit.

**Objective:** To develop a novel decision-making tool for physicians treating NSCLC patients on whether to administer immune checkpoint inhibitor (ICI) therapy alone or in combination with chemotherapy.

**Design, setting, and participants:** This multicenter observational study includes patients from an ongoing clinical trial (PROPHETIC; NCT04056247). Patients were recruited from 13 different centers (total n=425; 58 patients were excluded) from June 2016 and June 2021. Plasma samples were obtained prior to treatment initiation, and deep proteomic profiling was conducted. PROphet® computational model for predicting clinical benefit (CB) probability at 12 months was developed based on the plasma proteomic profile. The model performance was validated in a blinded manner. Following validation, training and prediction was performed over the entire cohort using cross-validation methodology. The patients were divided into four groups based on their PD-L1 expression level combined with their CB probability, and the survival outcome was examined for each group. The data were analyzed from July to October 2022.

**Main outcome and measures:** Clinical benefit from ICI-based treatment, overall survival (OS) and progression-free survival (PFS).

**Results:** The model displayed strong predictive capability with an AUC of 0.78 (p-value = 5.00e-05), outperforming a PD-L1-based predictive model (AUC = 0.62; p-value 2.76e-01), and exhibited a significant difference in OS and PFS between patients with low and high CB probabilities. When combining CB probability with PD-L1 expression levels, four patient subgroups were identified; (i) patients with PD-L1≥50% and a negative PROphet result who significantly benefit from ICI-chemotherapy combination therapy compared to ICI monotherapy; (ii) patients with PD-L1≥50% and a positive PROphet result who benefit similarly from either treatment modalities; (iii) patients with PD-L1<50% and a negative PROphet result who do not benefit from either treatment modalities; (iv) patients with PD-L1<50% and a positive PROphet score who benefit from combination therapy.

**Conclusions and relevance:** The PROphet® model displayed good performance for prediction of CB at 12 months based on a plasma sample obtained prior to treatment. Our findings further demonstrate a potential clinical utility for informing treatment decisions for NSCLC patients treated with ICIs by adding resolution to the PD-L1 biomarker currently used to guide treatment selection, thereby enabling to select the most suitable treatment modality for each patient.

## Introduction

ICI-based therapies are standard of care for NSCLC patients^1-3^. According to current guidelines for driver mutation-negative NSCLC, patients with PD-L1 expression ≥50% are treated with first-line immune checkpoint inhibitor (ICI) monotherapy or combination ICI-chemotherapy. For patients with PD-L1 expression <50%, combination ICI-chemotherapy is the preferred choice^4^. However, clinical evidence demonstrates limitations of the PD-L1 biomarker in predicting benefit from ICI-based therapy^5-7^. Predictive biomarkers with greater accuracy are therefore needed for guiding treatment choices.

Here, we developed PROphet®, a novel and robust machine learning (ML)-based model that analyzes proteomic profiles in pre-treatment blood plasma to predict benefit from ICI-based therapy and optimize treatment selection.

## Methods

### Patient cohort

Blood plasma samples and clinical data were collected from advanced stage NSCLC patients within the framework of the PROPHETIC clinical study (NCT04056247). All clinical sites received IRB approval for the study protocol and all patients provided written informed consent. Samples analyzed in this study were collected from 425 patients at 13 medical centers in Israel, Germany and USA between June 2016 and June 2021. Of the 425 enrolled patients, 58 were excluded due to technical or clinical reasons, resulting in 367 patients in the analyzed cohort (eFigure 1). Clinical data were collected for each patient. Patients displaying progression-free survival (PFS) before 12-months time point were classified as ‘no clinical benefit’ (NCB) patients. All other patients were classified as ‘clinical benefit’ (CB) patients.

### Proteomic-based computational model

Proteomic profiling of plasma samples was performed using the SomaScan^®^ Assay v4.1 (SomaLogic, Boulder, CO). The PROphet^®^ model was developed on the development set (n=254) and was tested in a blinded manner on the independent validation set (n=85), while the rest of the samples were left out from the prediction analysis as they were included in a previous round of training and validation. The division was done randomly while maintaining an equal distribution of main clinical features between the two sets (eMethods and eTable 1 in the Supplement). A set of 388 proteins that displayed differential plasma level distributions in CB and NCB populations was identified using Kolmogorov-Smirnov statistical test following 80 iterations of training and test sets selection (eMethods and eFigure2 in the Supplement). Such proteins, termed resistance associated proteins (RAPs), serve as potential indicators of CB in the following manner: for a given patient, a machine learning-based model infers a CB or NCB prediction for each one of the 388 RAPs based on XGBoost algorithm; the sum of all predictions in a given patient, called RAP score, reflects the patient’s likelihood of benefiting from treatment (patients displaying numerous CB predictions are more likely to benefit, and vice versa). To generate the output of the PROphet^®^ model, termed as the clinical benefit probability (CB probability), RAP scores were linearly scaled to values between 0 and 1.

## Results

Patient clinical parameters are presented in Table 1. The median age was 65 years with a predominance of male patients (approximately 2:1). The majority of the patients (78%) had non-squamous cell carcinoma, in agreement with expected proportions. Most of the patients had ECOG performance status of 0-1 (95%). Patients were either treated with ICI-chemotherapy combinations (56%) or ICI monotherapy (44%). There was an approximately equal distribution of patients with PD-L1<1%, PD-L1 1-49% and PD-L1≥50% tumors. Twenty five percent of the patients achieved CB at 12 months.

**Table 1:**
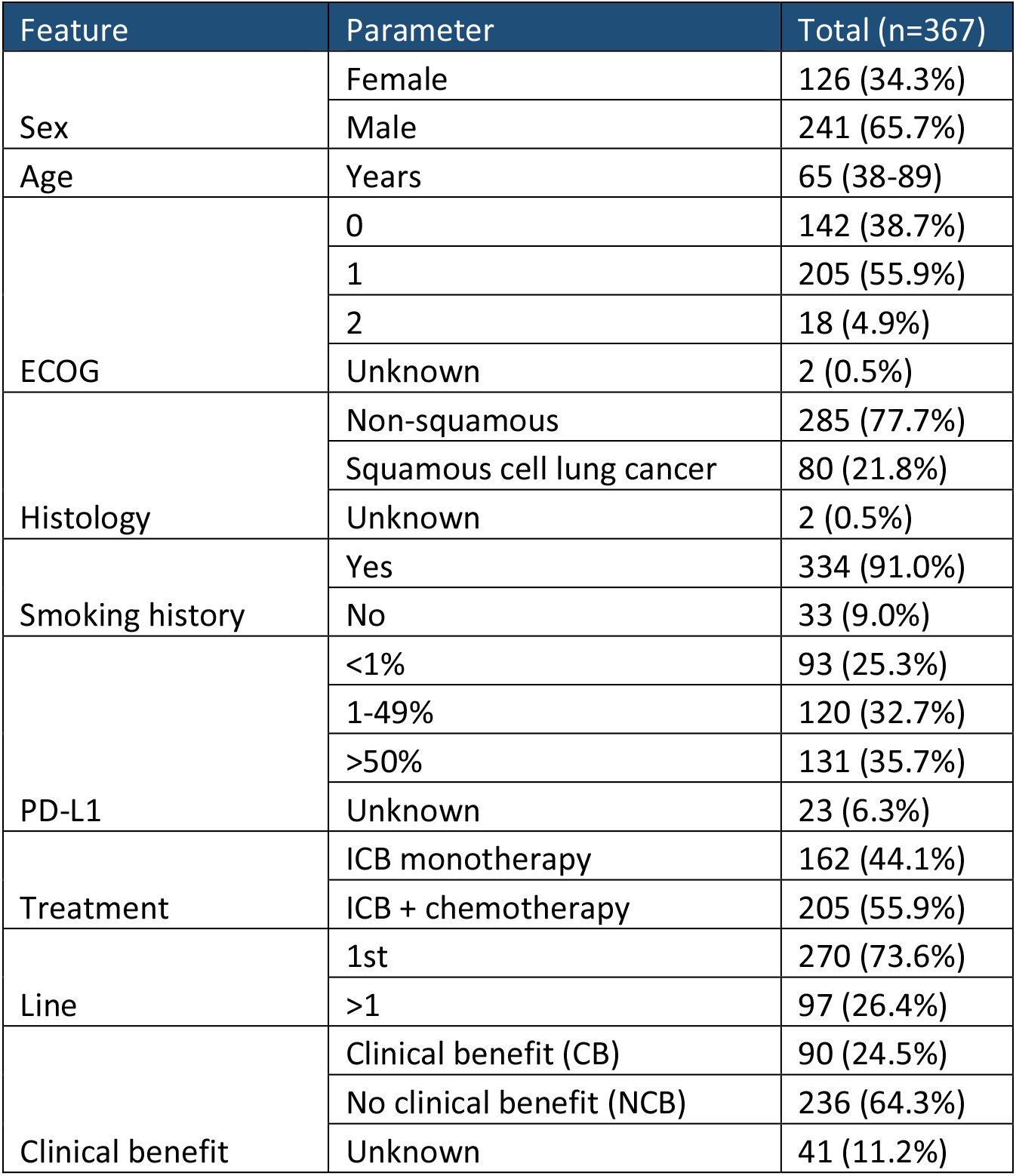
Basic clinical parameters. For each parameter, except for age, the total is presented, and the percentage is presented in brackets. Age is presented as median and the range in brackets.

As PD-L1 expression is a companion diagnostic test that guides treatment decision in NSCLC patients without driver mutations, we began by evaluating the predictive performance of a model based on the PD-L1 biomarker. The PD-L1 based model displayed area under the curve (AUC) of the receiver operating characteristic (ROC) plot of 0.55 (Figure 1A) and hazard ratio (HR) of 0.95 (Figure 1B). Next, we developed a clinical model based on PD-L1 and additional parameters previously shown to associate with clinical benefit, namely ECOG^8^, sex^9^ and line of treatment^10^. The clinical model displayed a minor improvement over the PD-L1-based model, with an AUC of 0.62 (Figure 1A-B). Aiming to develop a more robust predictive model, we developed an ML-based model, termed PROphet^®^. The output (CB probability) is based on the sum of predictions from a large collection of proteomic biomarkers associated with therapeutic benefit (eMethods in the supplement). The PROphet^®^ model displayed superior predictive performance in comparison to the PD-L1-based and clinical models (AUC=0.78; Figure 1A). In addition, a log-rank test demonstrated that patients with high CB probability achieved significantly longer PFS and OS than patients with low CB probability (Fig. 1B-D, HR = 0.41 and 0.38, respectively), further outperforming PD-L1 based predictions. Lastly, linear regression analysis demonstrated a high goodness of fit between predicted CB probability and observed CB rate (R^2^ = 0.94; Fig. 1E). Notably, actual NCB patients clustered at the lower range of predicted CB probabilities, indicating that the model has a high predictive power (Fig. 2F). This finding was further strengthened by an enrichment analysis (2D enrichment test; False discovery rate < 0.05; eFigure 3).

**Figure 1:**
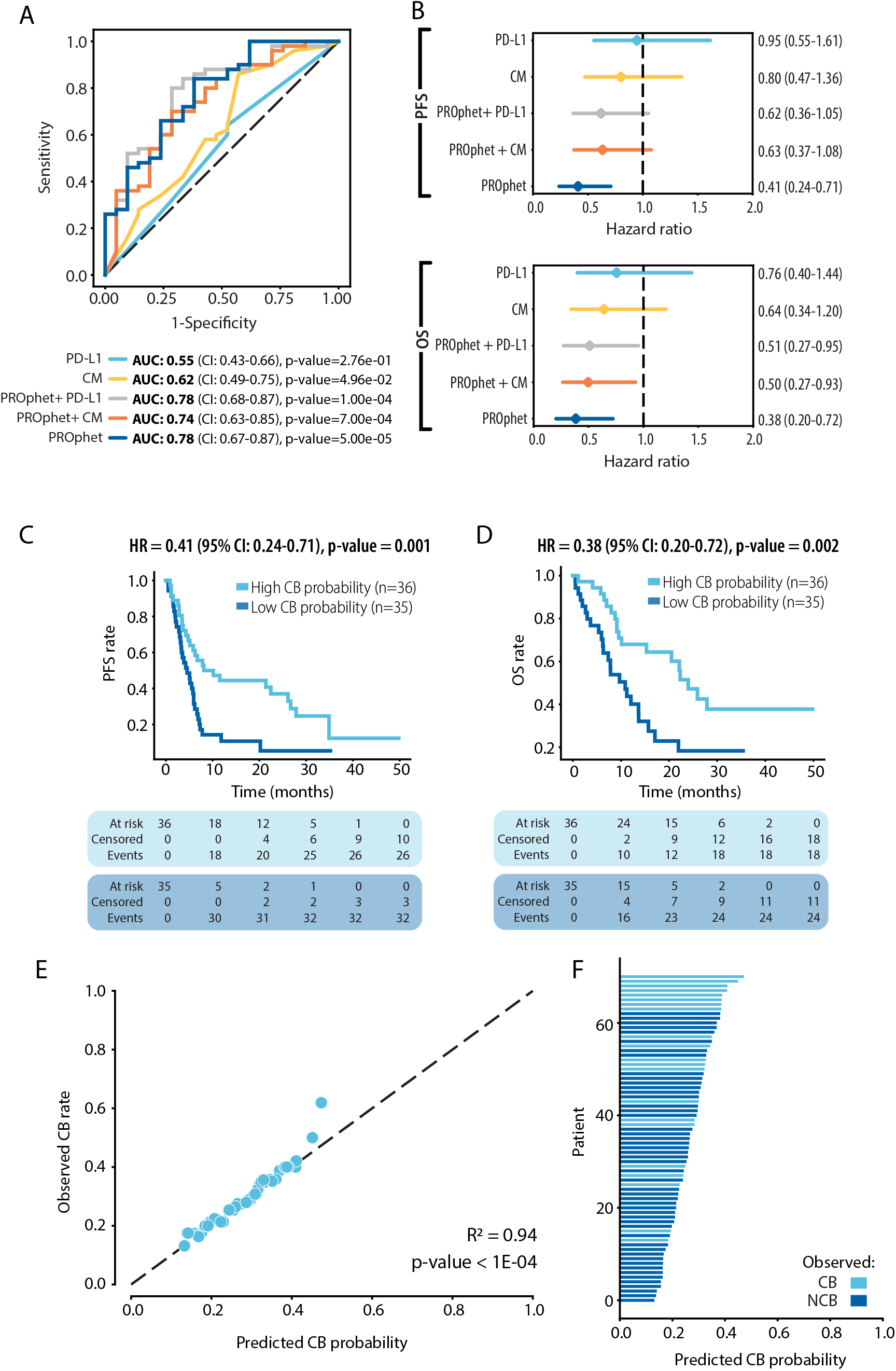
Performance of the PROphet^®^ predictive model. A. ROC plots of the five models. Predictive performance was compared across five models: PROphet^®^ model; PD-L1-based model (PD-L1); Clinical model (CM); Integrated PROphet^®^ + PD-L1; Integrated PROphet^®^ + CM. (B). The AUC is indicated. The dashed line indicates AUC=0.5. CI, confidence interval. B. Forest plot comparing the five models. Cox regression analysis based on OS (top) and PFS (bottom) data. C. and D. OS (C) and PFS (D) analysis of patients stratified to high and low CB probability groups. The median CB probability was used as the stratification threshold. HR, hazard ratio. CI, confidence interval. E. Predicted CB probability as a function of observed CB rate. Each dot represents a patient. The observed CB rate for each predicted CB probability datapoint refers to the proportion of observed CB patients within a patient group assigned CB probability ±0.15. X=Y is indicated by a black line. The goodness of fit is indicated. F. Bar plot showing predicted clinical benefit (CB) probabilities sorted from lowest.

**Figure 2:**
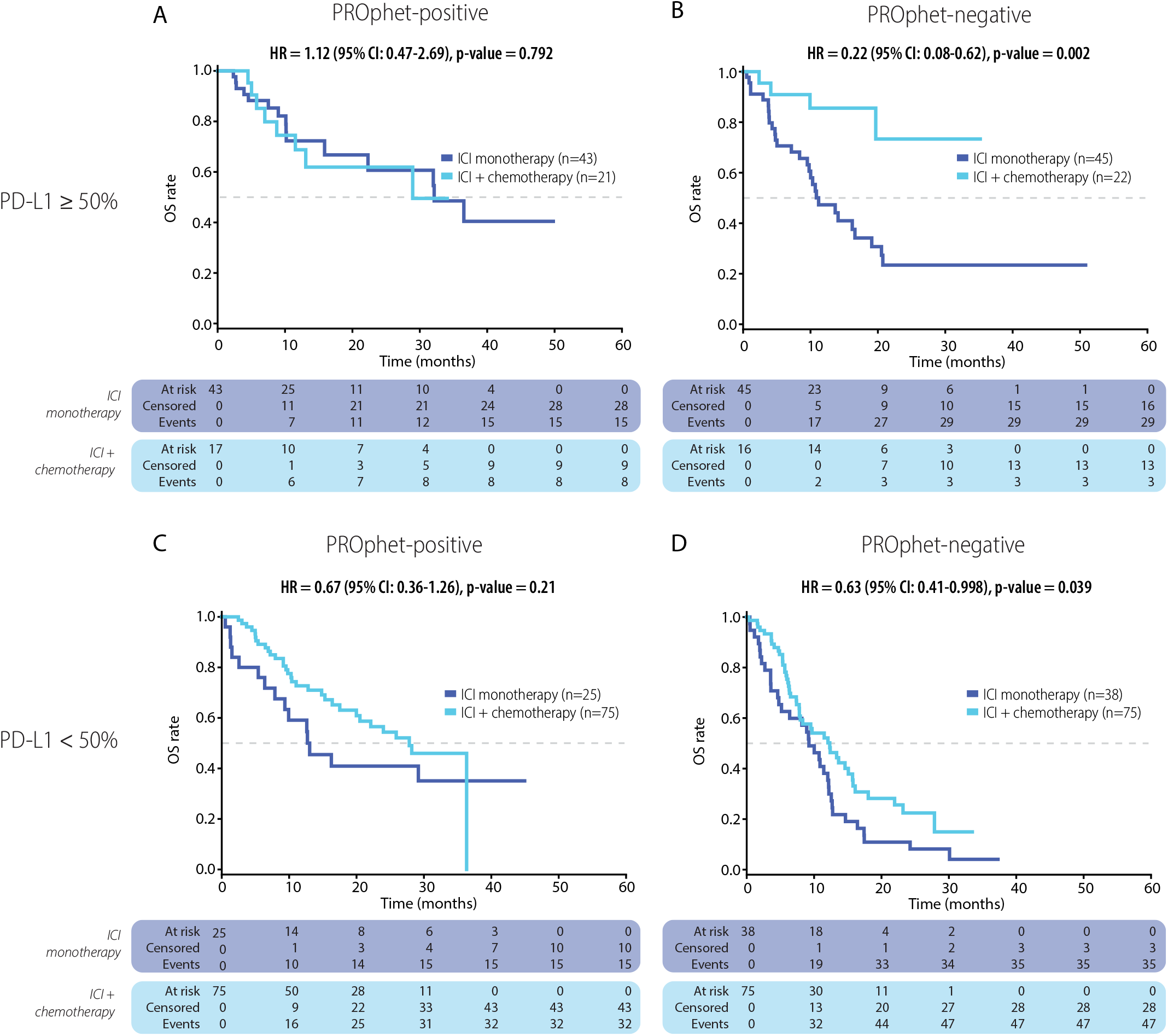
The PROphet^®^ model predicts differential overall survival outcomes when combined with PD-L1 expression level. A. and B. PD-L1≥50% patients were stratified to PROphet^®^ positive (A) and PROphet^®^ negative (B) groups. C. and D. PD-L1<50% patients were stratified to PROphet^®^ positive (C) and PROphet^®^ negative (D) groups. OS was evaluated in patients treated with ICI monotherapy vs combination ICI-chemotherapy. Dashed line indicates median survival.

Next, we tested the ability of the PROphet^®^ model to forecast survival outcomes in PD-L1≥50% patients receiving ICI monotherapy or combination ICI-chemotherapy. Patients were classified into PROphet^®^ negative and positive groups using the median CB probability as a threshold. In the PROphet^®^-positive group, patients receiving ICI monotherapy or combination therapy fared similarly well (Figure 2A and eFigure 4A in the supplement). This implies that such patients are suitable candidates for monotherapy and may be spared the more toxic ICI-chemotherapy combination. In contrast, in the PROphet^®^-negative group, OS and PFS were significantly longer in patients receiving ICI-chemotherapy in comparison to ICI monotherapy (Fig. 2B and eFigure 4A in the supplement). Median OS was not reached versus 11.20 months (combination vs monotherapy; HR=0.22; p=0.002) and median PFS was 14.29 vs. 5.52 months (combination vs. monotherapy; HR=0.44; p=0.019). This suggests that PD-L1≥50% patients with a PROphet^®^-negative score should consider combination ICI-chemotherapy despite high PD-L1 levels.

Finally, we asked whether the model could provide insights for managing patients with PD-L1 <50%. In this analysis, patients in the PROphet^®^-positive group displayed an OS benefit when treated with ICI-chemotherapy combination in comparison to patients receiving monotherapy, although statistical significance was not reached (Figure 2C). Median OS was 27.83 months for ICI-chemotherapy vs 13.04 months for ICI monotherapy. This result is in line with current guidelines recommending ICI-chemotherapy for patients with PD-L1<50%. In contrast, PROphet^®^-negative patients displayed poor outcomes when treated with either of the two treatment modalities, with a median OS of 9.32 and 12.32 months for monotherapy and ICI-chemotherapy, respectively (Fig. 2D). These findings suggest that treatment types other than the standard of care may be considered for these patients.

## Discussion

Here we describe a novel a tool for supporting treatment decision for NSCLC patients receiving anti-PD-(L)1-based therapy. The PROphet^®^ model provides two main clinical utilities. First, it successfully predicts therapeutic benefit at 12 months, displaying superior predictive capabilities over PD-L1 based models. Second, when used in combination with PD-L1 testing, the model helps in determining whether a patient should receive ICI alone or an ICI-chemotherapy combination. Thus far, this tool can potentially improve overall rate by guiding patients with PROphet^®^ negative result to alternative treatment modalities. Notably, tumor PD-L1 expression and tumor mutational burden (TMB) are the most prominent biomarkers for predicting clinical benefit from ICI-based therapy to date. While these biomarkers require tumor tissues, which is sometimes not available, PROphet^®^ requires a blood test, which simplifies the procedure.

Our model analyzes a large set of proteomic biomarkers, termed RAPs, that collectively provide a robust prediction of treatment benefit. Using supervised computational models to analyze omic-level data, we and others have previously identified blood-based signatures predictive of ICI outcomes^11-14^. However, successful validation can be challenging due to several intrinsic limitations, including overfitting and inter- and intra-patient heterogeneity, while large sample sizes are necessary to achieve acceptable model performance. The PROphet^®^ model handles such challenges due to its reliance on hundreds of proteomic predictors. Assuming that most RAPs are indeed associated with CB, those RAPs that yield less reliable predictions have a minimal impact on the final output.

This study has several limitations. First, the analyzed cohort includes patients receiving different treatment modalities, as well as patients with tumors of different histological types. Second, the cohort does not include a control group, and the patients were not randomized among the therapeutic strategies. Therefore, interpretation is limited. Third, while we tested the model in an independent validation set, this patient set was relatively small. Validation in a larger cohort is required.

## Conclusions

Altogether, this study shows that using proteomic analysis of a pre-treatment plasma sample, the PROphet^®^ model, when combined with PD-L1 test, stratifies the patients into four subgroups, providing additional resolution to the PD-L1 biomarker currently used to guide treatment selection.

## Data Availability

Data produced in the present study are available upon reasonable request to the authors

## Supplementary figure legend

**eFigure 1:** Patient exclusion. A cohort of 425 patients was assembled. Following patient exclusion, 339 patients remained in the analysis. Reasons for exclusion are indicated.

**eFigure 2**: A. Development of the PROphet^®^ prediction model. A cohort of advanced stage NSCLC patients receiving ICI-based therapy was assembled. Pre-treatment blood samples were obtained, and plasma proteomes were profiled using SomaScan^®^ technology. Clinical benefit (CB) was assessed at 12 months after starting treatment, and patients were followed up for 2 years. A predictive model for CB was developed as follows: Proteins displaying differential plasma levels in CB and NCB patient populations were selected for model training using a statistical test. Such proteins are collectively termed Resistance Associated Proteins (RAPs). A predictive model for CB was developed per RAP using a machine learning algorithm. CB predictions inferred from each RAP were summed up to yield a RAP score per patient. RAP scores were linearly scaled to values between 0 and 1, enabling the conversion of a given patient’s RAP score into a CB probability. B. Development and validation of the RAP model. The cohort was divided into development and validation sets (75% and 25%, respectively). The development set was randomly divided into train and test sets (75% and 25%, respectively). The train set was used for RAP selection followed by model training resulting in a predictive model per RAP. Clinical benefit (CB) predictions were then generated per RAP for each patient in the test set. CB predictions from all selected RAPs were summed up to yield a RAP score per patient in the test set. The process was repeated 80 times, each time with a random division of development set patients into train and test sets. RAP scores were averaged per patient in the development set and linearly scaled. Model output is CB probability (a value between 0 and 1). The model was then locked and tested on the independent validation set.

**eFigure 3**: Enrichment analysis for CB probabilities and observed CB rates at each time point. The enrichment analysis was done using 2D-enrichment test. The X-axis indicates the enrichment score for predicted CB probability. The Y-axis indicates the enrichment score for observed rates (as defined by the proportion of observed CB patients within a patient group assigned the CB probability ±0.15). The enrichment score is a value between 1 and −1. Positive and negative enrichment scores indicate enrichment in high and low CB probabilities, respectively and in high and low observed CB rates, respectively. The solid line indicates the X=Y line.

**eFigure 4:** The PROphet^®^ model predicts differential PFS outcomes when combined with PD-L1 expression level. A. and B. PD-L1≥50% patients were stratified to PROphet^®^ positive (A) and PROphet^®^ negative (B) groups. C. and D. PD-L1<50% patients were stratified to PROphet positive (C) and PROphet^®^ negative (D) groups. PFS was evaluated in patients treated with ICI monotherapy vs combination ICI-chemotherapy. Dashed line indicates median PFS.

## References

1. Borghaei H, Paz-Ares L, Horn L, et al. Nivolumab versus Docetaxel in Advanced Nonsquamous Non-Small-Cell Lung Cancer. N Engl J Med. Oct 22 2015;373(17):1627–39. doi:10.1056/NEJMoa1507643

2. Reck M, Rodríguez-Abreu D, Robinson AG, et al. Pembrolizumab versus Chemotherapy for PD-L1-Positive Non-Small-Cell Lung Cancer. N Engl J Med. 11 10 2016;375(19):1823–1833. doi:10.1056/NEJMoa1606774

3. Gaissmaier L, Christopoulos P. Immune Modulation in Lung Cancer: Current Concepts and Future Strategies. Respiration. Dec 08 2020:1–27. doi:10.1159/000510385

4. Hanna NH, Schneider BJ, Temin S, et al. Therapy for Stage IV Non-Small-Cell Lung Cancer Without Driver Alterations: ASCO and OH (CCO) Joint Guideline Update. J Clin Oncol. 05 10 2020;38(14):1608–1632. doi:10.1200/JCO.19.03022

5. Scognamiglio G, De Chiara A, Di Bonito M, et al. Variability in Immunohistochemical Detection of Programmed Death Ligand 1 (PD-L1) in Cancer Tissue Types. Int J Mol Sci. May 21 2016;17(5) doi:10.3390/ijms17050790

6. McLaughlin J, Han G, Schalper KA, et al. Quantitative Assessment of the Heterogeneity of PD-L1 Expression in Non-Small-Cell Lung Cancer. JAMA Oncol. Jan 2016;2(1):46–54. doi:10.1001/jamaoncol.2015.3638

7. Hofman P. PD-L1 immunohistochemistry for non-small cell lung carcinoma: which strategy should be adopted? Expert Rev Mol Diagn. 12 2017;17(12):1097–1108. doi:10.1080/14737159.2017.1398083

8. Dall’Olio FG, Maggio I, Massucci M, Mollica V, Fragomeno B, Ardizzoni A. ECOG performance status ≥2 as a prognostic factor in patients with advanced non small cell lung cancer treated with immune checkpoint inhibitors-A systematic review and meta-analysis of real world data. Lung Cancer. 07 2020;145:95–104. doi:10.1016/j.lungcan.2020.04.027

9. Conforti F, Pala L, Bagnardi V, et al. Cancer immunotherapy efficacy and patients’ sex: a systematic review and meta-analysis. Lancet Oncol. 06 2018;19(6):737–746. doi:10.1016/S1470-2045(18)30261-4

10. Lang D, Huemer F, Rinnerthaler G, et al. Therapy Line and Associated Predictors of Response to PD-1/PD-L1-Inhibitor Monotherapy in Advanced Non-small-Cell Lung Cancer: A Retrospective Bi-centric Cohort Study. Target Oncol. 12 2019;14(6):707–717. doi:10.1007/s11523-019-00679-9

11. Harel M, Lahav C, Jacob E, et al. Longitudinal plasma proteomic profiling of patients with non-small cell lung cancer undergoing immune checkpoint blockade. J Immunother Cancer. Jun 2022;10(6) doi:10.1136/jitc-2022-004582

12. Muller M, Hummelink K, Hurkmans DP, et al. A Serum Protein Classifier Identifying Patients with Advanced Non-Small Cell Lung Cancer Who Derive Clinical Benefit from Treatment with Immune Checkpoint Inhibitors. Clin Cancer Res. Oct 01 2020;26(19):5188–5197. doi:10.1158/1078-0432.CCR-20-0538

13. Nabet BY, Esfahani MS, Moding EJ, et al. Noninvasive Early Identification of Therapeutic Benefit from Immune Checkpoint Inhibition. Cell. Oct 15 2020;183(2):363-376.e13. doi:10.1016/j.cell.2020.09.001

14. Rich P, Mitchell RB, Schaefer E, et al. Real-world performance of blood-based proteomic profiling in first-line immunotherapy treatment in advanced stage non-small cell lung cancer. J Immunother Cancer. Oct 2021;9(10) doi:10.1136/jitc-2021-002989

